# Addressing the Global Diagnostics Gap for Childhood Leukemias: A Global, Multisite Type 2 Hybrid Validation Study of Nanopore-based Adaptive Sampling Whole Genome Sequencing

**DOI:** 10.64898/2026.05.19.26353434

**Authors:** Thomas B. Alexander, Rubina Islam, Javeria Aijaz, Nancy Bolous, Karlijn Cammel, Julie Geyer, Shannon Gray, Shakir Hussain, Sadia Imran, Saba Jamal, Srijita Kar, Dona Kanavy, Neelum Mansoor, Mayur Parihar, Vaskar Saha, Bastiaan Tops, David Wilkins, Karen Weck, Gang Wu, Liang Zhou, Lennart Kester, Jeremy Wang, Nickhill Bhakta

## Abstract

**Background:** Modern therapy for childhood and adolescent leukemia requires accurate risk classification of genomic subtype. Although short-read next-generation sequencing (NGS)-based approaches provide comprehensive clinical diagnostics in limited, highly resourced settings, they remain expensive, slow, and inaccessible to most children worldwide. Transformative approaches are needed to improve diagnostic classification for leukemia globally.

**Methods:** We simultaneously continued to develop an analytical pipeline NASVar (Nanopore variant calling for adaptive sampling), and conducted a multicenter, type-two hybrid clinical validation study of an Oxford Nanopore Technologies (ONT) adaptive-sampling whole-genome sequencing (asWGS) assay across hospitals with varying diagnostic resources. In preparation for implementation, a global panel developed a leukemia-based standardized gene set and consensus laboratory-developed test (LDT) validation guidelines. Measures of assay effectiveness compared to both conventional and orthogonal NGS methods, where available, were simultaneously collected with data to measure the implementation outcomes of feasibility, fidelity, appropriateness, and cost.

**Results:** All four centers successfully completed the LDT validation, with minimal adaptations required for regulatory compliance. A total of 457 specimens were sequenced (331 B-ALL, 83 AML, 43 T-ALL). For the 210 B-ALL cases with locally resolved genomic subtypes defined by DNA alterations, asWGS was 100% concordant (210/210). Cases locally defined as B-other were resolved via asWGS with disease-defining DNA alterations in 47% (49/105) of cases. An additional 41% (43/105) of locally defined B-other cases were classified by incorporation of DNA methylation, and all 16 B-ALL patient-derived xenograft controls were correct, for a total of 96% (318/331) of all B-ALL cases in the cohort resolved with single assay asWGS. For AML, 97% (56/58) of cases with locally resolved genomic subtypes were identified by automated asWGS analysis, while an additional two cases were identified after targeted manual review. At Indus Hospital in Pakistan, the B-ALL and AML diagnostic genomic subtype yield increased from 28% with local standard of care diagnostic testing, to 84% with asWGS. The cost of reagents and consumables in the United States, assuming pooled three-plexing, was $343/sample. Based on the combined hybrid validation results, all centers are independently preparing for clinical return of results.

**Conclusions:** ONT asWGS was successfully validated as a clinical assay in four diverse hospital settings. As a single, multi-omic platform that delivers value across the continuum of high-resource to resource-limited contexts, the approach offers a disruptive solution to address the global equity gap in cancer diagnostics.

## Introduction

Over the past sixty-five years, advances in clinical therapeutics, supportive care, and risk stratification have improved 5-year net survival for children who are diagnosed with leukemia from near universal mortality to >85% in high-income countries.(1,2) More recently, advances in immunotherapy have led to revolutionary changes in the upfront and relapse management of childhood B-cell acute lymphoblastic leukemias (B-ALL) (3,4) These successes have led to integration of complex therapeutic treatments into upfront clinical practice guidelines and justified the design of initiatives such as the World Health Organization and St. Jude Children’s Research Hospital’s Global Platform for Access to Childhood Cancer Medicines (GPACCM).(5–7)

As treatments continue to improve, genomic classification of acute leukemias has become integral when determining optimal management.(8) Next-generation sequencing (NGS) approaches, such as whole-genome sequencing (WGS), whole-transcriptome sequencing (WTS), and methylation, that provide orthogonal and unique diagnostic information are now recommended in comprehensive management guidelines and increasingly required in clinical trials.(5,6) Yet, despite recent advances in evidence-based modalities to treat acute leukemias, a lack of innovation to improve universal access to clinical genomics remains. Due to technical demands, high costs, long turnaround times, and analytical challenges, only a small number of centers, predominantly in high-income countries, routinely use NGS for clinical care. (9–12). In addition, for the 90% of children living in low- and middle-income countries, broad access to all molecular modalities remains firmly out of reach.(13–15) Thus, equitable diagnostic solutions that overcome observed access-inhibiting infrastructure and logistical challenges and are context-appropriate across all income-level settings are needed.

Adaptive sampling whole-genome sequencing (asWGS), a real-time, software-driven method using Oxford Nanopore Technologies (ONT) devices, is a newer sequencing method that has the potential to address many of the barriers described that limit the standard clinical use of NGS.(16) Recently, our group demonstrated that asWGS can identify biologically relevant structural variants, aneuploid, internal duplications and deletions, analytically targeted single-nucleotide variants, copy number variants of different sizes, and methylation, with all information derived from a singular assay.(17–20) (**Figure 1**) However, these works were completed within a single research lab as a proof-of-concept study. To evaluate real-world clinical effectiveness and strategies for the rapid scaling of this assay, we conducted a multicenter, type two hybrid effectiveness-implementation clinical diagnostic validation study among four hospitals with varying conventional diagnostic resources with a goal of identifying a novel strategy to address the childhood leukemia diagnostics access gap.

**Figure 1:**
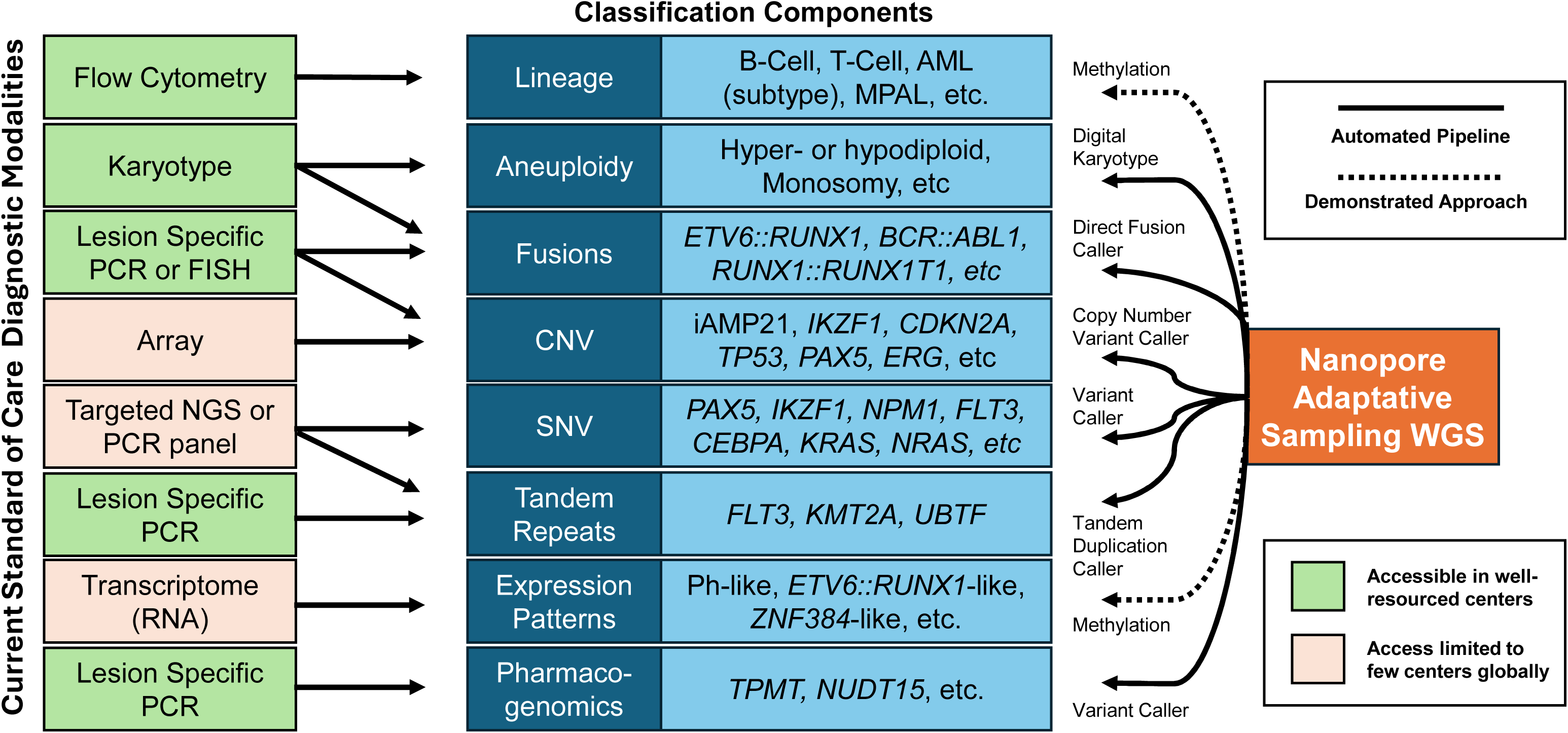
Comparison of Conventional Pathology Testing Strategy Outcomes versus Nanopore Adaptive Whole Genome Sequencing for the Diagnosis of Pediatric Acute Leukemias

## Methods

### Study Design Overview

We used a type two hybrid effectiveness-implementation design to study the clinical validation of an asWGS-based leukemia assay for a diagnostic assay at four hospitals globally. Effectiveness of the assay was measured by comparing genomic subtypes of ALL and AML between asWGS and locally available standard care pathology modalities. Simultaneously, we also measured feasibility, fidelity, appropriateness, and cost as implementation outcomes. Validation of the assay was conducted in a quasi-independent manner. Although all centers agreed to use a common laboratory-developed test (LDT) framework, each had full latitude to adapt the tool based on local regulatory requirements.

### Pre-implementation Strategies

To support validation, two working groups with international representation from researchers, pathologists, and clinicians were formed to create a (1) standardized gene set (Supp. Table 1) and a (2) common LDT framework. Working group members met virtually twice and reached a consensus. The analytic and computational basis of the assay was previously published.(17)

### Participating Laboratories and Effectiveness Evaluation

Four hospitals from among the working groups above, with diverse clinical, research, and income characteristics, were immediately ready to participate in the validation study cohort: The University of North Carolina at Chapel Hill (UNC), United States; Tata Medical Center, Kolkata (TMC-K), India; Indus Hospital and Health Network (IHHN), Pakistan; Princess Máxima Center for Pediatric Oncology (PMC), Netherlands. UNC has access to comprehensive conventional pathology assays but does not routinely conduct NGS for all new diagnosis patients. TMC-K uses an advanced conventional diagnostic algorithm for all new diagnosis leukemias and has access to short-read gene panels and WTS testing on a limited basis. PMC has comprehensive conventional and short-read NGS for all cases (WGS and WTS), allowing for orthogonal ground-truth effectiveness comparisons. IHHN performs a limited conventional diagnostic panel, consisting of flow cytometry, karyotype and FISH probes (B-ALL: *ETV6::RUNX1*, *TCF3::PBX1*, *KMT2A*r; AML: *RUNX1::RUNX1T1*, *KMT2A*r), and has no access to NGS. To provide an assessment of clinical utility in a setting with limited diagnostic capacity, an unbiased, nested cohort of 74 sequentially diagnosed patients from the IHHN biobank was constructed within their LDT sample set.

### Specimen preparation and sequencing

Bone marrow or peripheral blood samples were collected as clinical discard or for explicit biobanking for research. Samples included fresh specimens, specimens stored as isolated DNA, and cell suspensions preserved in Zymo DNA/RNA Shield (Zymo Research Corporation). Standard DNA extraction, library preparation, and sequencing SOPs were reviewed and adopted by each participating center. To optimize asDNA sequencing, our protocol includes a mechanical shearing step that generally targets DNA fragments in the range of 5000 base pairs. As an external reference set, each site also sequenced the same five patient-derived xenograft specimens from St. Jude Children’s Research Hospital PROPEL Data Portal (https://propel.stjude.cloud).

### Data Analysis

Basecalling was performed using Dorado (ONT, model v5.0.0) in high- or super-accurate mode. Reads were aligned to the T2T CHM13v2.0 reference genome using minimap2.(21). Digital karyotyping and variant analysis was performed using an automated analysis pipeline (NASVar) from aligned BAM files as previously described with additional improvements such as inclusion of minor allele frequencies to support aneuploidy calls for digital karyotypes, calling of single-sided fusions, and additional automation of specific CNV and SNVs.(17) A list of NASVar automated calls derived from the analytical pipeline modules is provided in Supplemental Table 2. Methylation results were generated using tools developed by PMC.(20)

### Effectiveness Outcomes

Accuracy, precision, and replicability were all evaluated at the cohort level, and concordance data were summarized with confusion matrices. A NASVar report was automatically generated for each sample in the cohort. Results were reviewed by the local clinical lead and compared to available orthogonal clinical testing. Each B-ALL and AML sample was categorized into a genomic subtype according to published clinical practice guidelines, except for B-ALL subtypes defined by GEP classification (*PAX5*alt, Ph-like without canonical fusion, *ZNF384*r-like, *ETV6::RUNX1*-like, and *KMT2A*r-like). GEP-requiring classes were categorized as “B-Other” for the primary analysis to allow for comparable evaluation of NASVar versus real-world clinical testing. In a secondary analysis of B-Other samples, asWGS derived methylation results were integrated with NASVar findings to provide further characterization and comparison to short-read WTS GEP classification calls where available. For T-ALL, which lacks a uniform, clinically relevant genomic categorization, exemplar categories demonstrating clinical utility were selected based on common subtypes. For samples where methylation and NASVar results were discordant, fastq files were manually reviewed for DNA alterations in a targeted manner based on methylation classification. All discordant or ambiguous results between clinical testing and asWGS were reviewed centrally by the study leadership team.

### Implementation Outcomes

Feasibility was defined by the ability for all centers to complete the LDT framework. Fidelity was measured based on adaptations to the framework, gene set, and SOPs. Adaptations were analyzed using FRAME-IS.(22) Appropriateness domains of suitability, fit, and ultimate relevance were evaluated based on whether centers intend to implement the assay as a routine clinical diagnostic following local LDT completion and analytic assessment.

To evaluate costs, a microcosting analysis of reagents and consumables for DNA extraction, library preparation, quality control, and sequencing was completed based on a previously developed framework.(23) Activity-based costing was excluded as a sunk cost since the ONT assay is redundant with existing conventional pathology methods. An upper-limit willingness-to-pay threshold of $400/sample was qualitatively established prior to study initiation together with working group members from seven centers located in World Bank low- and middle-income settings. The impact of resequencing to achieve >20x on-target coverage to identify a canonical DNA alteration on cost thresholds was incorporated as a one-way sensitivity analysis. Additional details available in the Supplement.

## Findings

We performed asWGS on 457 specimens at four sites: 117 (57 previously published) (17) at UNC in the United States, 112 at TMC-K in India, 129 at PMC in the Netherlands, and 99 at IHHN in Pakistan. Twenty specimens (5/site) represented patient derived xenografts. (**Figure 2**)

**Figure 2:**
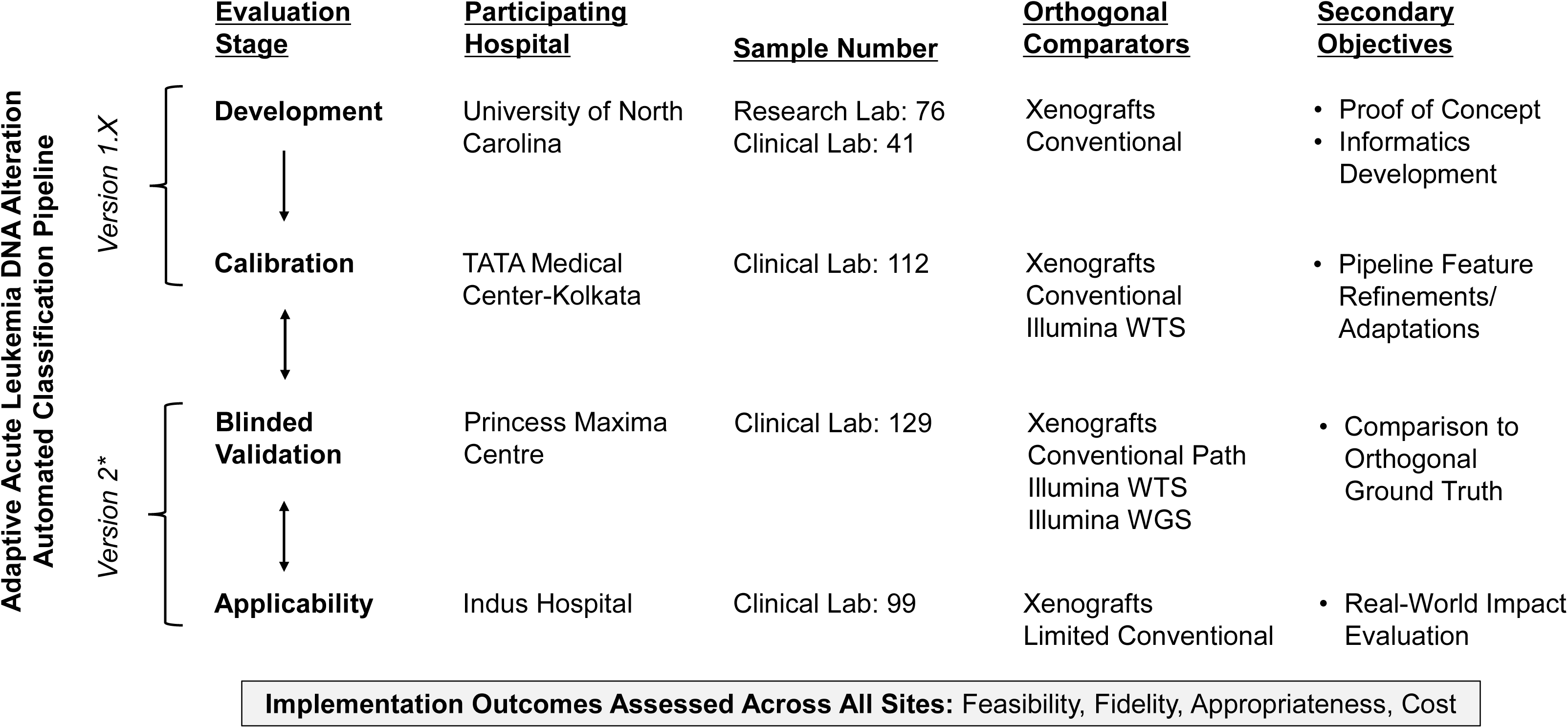
Study Design and Outcome Measures of the Type 2 Hybrid Effectiveness-Implementation Validation *Version 2 was subsequently rerun on all samples, inclusive of those first analyzed using version 1. All Results reported reflect Version 2

### Analytical Validation

#### Sequencing and Flow Cell Quality

Key sequencing quality metrics of on-target read length and mean coverage across target regions are presented in Supplementary Table 2. For all analytic samples, each center successfully achieved the expected on-target read length and the minimum coverage threshold for target regions. Per-sample coverage decreased with increasing multiplexing, as expected, and required repeat sequencing in certain instances at TMC-K and IHHN, with variation in flow cell output. (Supp. Table 3). Base-pair SNV accuracy compared to the reference genome was 99.1% across the cohort (Supp. Table 4). Seventeen samples required resequencing due to <20x on-target coverage without a driving alteration identified.

#### Limit of Detection

Dilution series were performed singleplex on a fusion-driven case and an aneuploid-driven case at two locations (UNC and PMC). (Supp. Table 5) The fusion breakpoints (*ETV6::RUNX1* or *TCF3::PBX1*) were detected to a dilution to 10% and detection was dependent on blast percentage and on-target depth. Aneuploid classification was correct at 50%, but lost sensitivity at dilution to 40% and was dependent on blast count alone; detection was not improved by increasing sequence depth.

#### Precision and Replicability

Five PROPEL xenograft specimens from the same cases (B-ALL hyperdiploid, *ETV6::RUNX1*, near haploid, and AML with *UBTF*-ITD) sequenced independently at each site were correctly classified. UNC and IHHN repeated sequencing on a minimum of 5 additional local samples, achieving 100% concordance in genomic subtype classification.

### Effectiveness Results

#### B-cell Acute Lymphoblastic Leukemia

67% (210/315) of B-ALL cases had an identified genomic subtype based on clinical testing for disease-defining alterations. All 210 were correctly identified using NASVar.(**Figure 3a**) This includes cases defined by common structural variants (*ETV6::RUNX1*, *TCF3::PBX1*, *KMT2A*r, and *BCR::ABL1),* complex or uncommon rearrangement (*IGH::DUX4*, *IGH::CEBPA*, *NUTM1*r, and *HLF*r), aneuploid (high-hyperdiploid cases, hypodiploid, iAMP21), and cases defined by a single point mutation, such as *PAX5* P80R. (**Figure 4**) Our automated digital karyotype provides discrete arm-level copy-number results, which is valuable for prognostic classification of groups based on specific chromosome deletions or gains. (**Figure 4**) (24).

**Figure 3:**
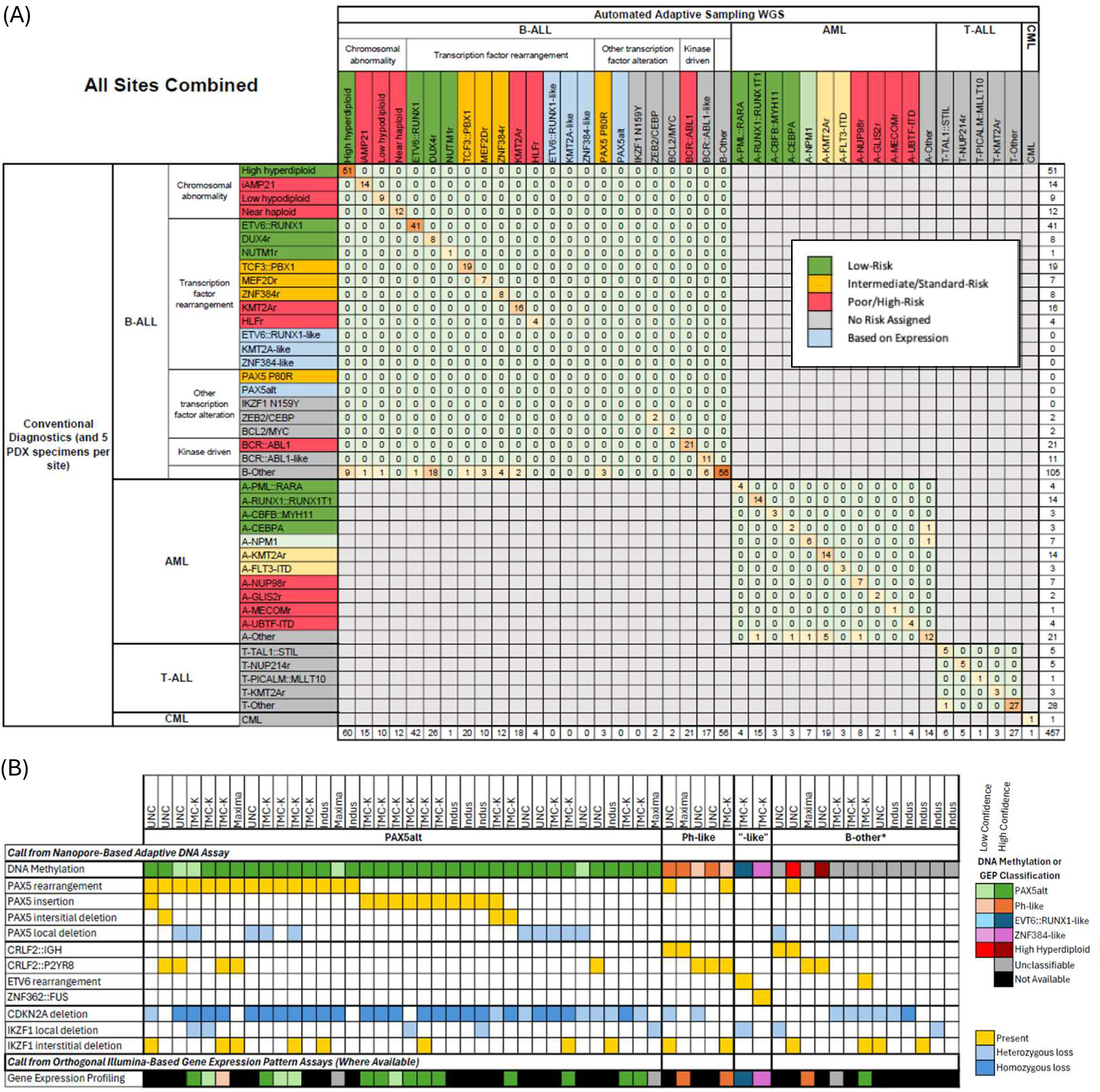
Effectiveness Outcomes Comparing DNA Alterations identified using NASVAar automated asWGS Compared to DNA Alterations Identified from Routine Clinical Testing. (a) Confusion Matrix Comparing Adaptative Sequencing Results Versus Final Conventional Pathology Classification at All Sites. (b) Oncoprint of B-ALL “Other” Subtypes Comparing DNA Methylation, NASVar Identified DNA Alterations and Short-Read Whole Transcriptomic Gene Expression Profiling *Risk categories based on NCCN guidelines for B-ALL and pediatric AML *A-*FLT3* is categorized if *FLT3*-ITD is identified in the absence of other subtype-defining lesions \**BCR-ABL1*-like for this table is defined by Ph-like defining fusions (i.e. we categorized *CRLF2*r as B-other for this table because *CRLF2*r can occur outside of Ph-like gene expression profiles) *A-other include a case of *CREBBP::KAT6A* and a cases of *CBFA2T3::RUNX1*, both driver fusions *patient-derived xenograft (PDX)

**Figure 4:**
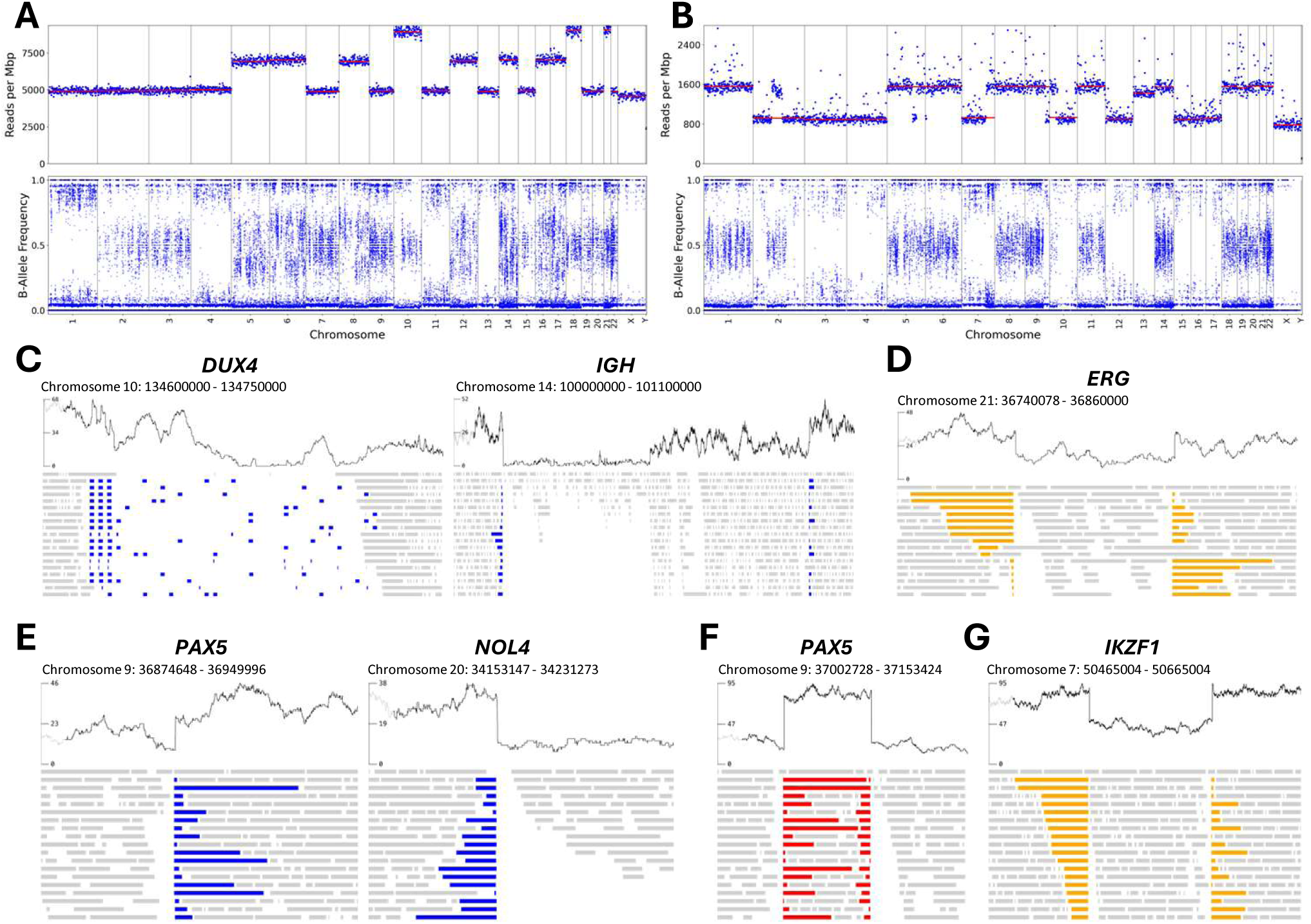
Examples of NASVar Output Among Clinically Interesting Leukemia Genomic Subtypes. (a) Hyperdiploid 60XY; +5, +6, +8, +10, +10, +12, +14, +16, +17, +18, +18, +21, +21, +X (b) Hypodiploid. 37X; -2, -3, -4, -7, -10q, -12, -15, -16, -17 (c) *DUX4-IGH* with concomitant (d) *ERG* deletion. TTCRC 35 (e) *PAX5-NOL4* (f) *PAX5* insertion (g) *IKZF1* intragenic deletion

Among the 33% (105/315) of cases without a disease-defining DNA alteration by clinical testing, 47% (49/105) were assigned to a specific B-ALL genomic subtype using asWGS by DNA alterations, and another 41% (43/105) were resolved using DNA methylation classification in addition to DNA alterations results from NASVar. Of the 43 B-ALL cases resolved with the incorporation of DNA methylation classification, 84% (36/43) were classified as *PAX5*alt by DNA methylation and 12% (5/41) were classified as Ph-like (**Figure 3b**). In the *PAX5*alt group, the NASVar variant calls matched the expected alterations in *PAX5*, with 15 cases of fusions, 10 focal insertions, 3 focal deletions, and 10 with copy loss. *CDKN2A* loss was observed in 86% (31/36) of *PAX5*alt cases. Of the remaining, the case classified as *ETV6::RUNX1*-like had an *ETV6* rearrangement found on NASVar, and another was classified as *ZNF384*r-like by DNA methylation with a *ZNF362::FUS* on NASVar. Four of the remaining unresolved cases contained *CRLF2*r, but without concordant DNA methylation support for Ph-like classification. Overall, 96% (317/331) of B-ALL cases sequenced across the four implementing centers were resolved by asWGS as a single assay.(**Figure 5**)

**Figure 5:**
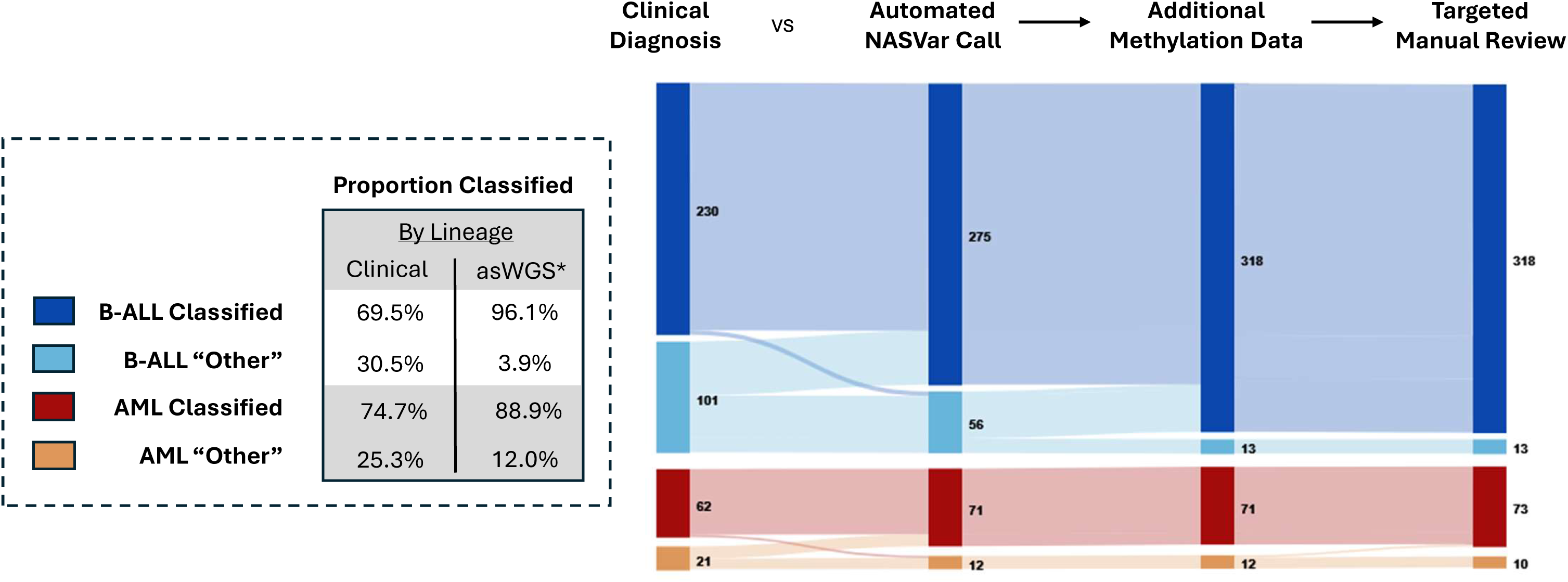
Sankey Diagram and Proportion of Samples with Classification Label based on Clinical Diagnosis and Integrated asWGS Calls Among All Samples in the Multisite Validation Cohort *Automated asWGS calls made using NASVar. In overall comparison of clinical vs asWGS, the results reflect the performance of integrating NASVar automated calls, methylation data and targeted manual review, applied in that hierarchical order. When NASVar DNA alterations were identified, no methylation data was utilized. When methylation data suggested a DNA alteration where none was identified based on NASVar, a targeted manual review of the region of interest was completed.

#### Acute Myeloid Leukemia and T-cell Acute Lymphoblastic Leukemia

Seventy-three percent (58/79) of AML cases were classified into genomic subtype by clinical testing. Ninety-seven percent (56/58) were identified using NASVar. Among the two discordant samples, NASVar output did not report one case with a 3-base pair deletion in the *CEBPA* bZIP region and another with an uncommon 24 bp insertion in *NPM1,* as these alteration types were not included in the version of NASVar used for this analysis (Supplement X). Methylation analysis correctly identified the discordant *CEPBA* sample with high confidence and suggested *NPM1* for the second case, but below the confidence threshold (score 0.7, threshold 0.9). On manual review of the asWGS fastq file, both alterations were identified. Of the 21 AML cases not classified by clinical testing, 52% (11/21) were identified by NASvar. For T-ALL, NASVar was 100% concordant with exemplar subtypes.

#### Value-add Where Clinical Classification is Limited

Of the 79 consecutive IHHN cases, 10 were T-ALL, 21 AML, and 43 B-ALL. Because B-ALL and AML rely heavily on genomic subtype for treatment decisions, we focused on these cases. The limited standard of care clinical testing in only Twenty-eight percent (18/64). Single assay asWGS classified 84% (54/64) of cases, tripling the number of cases with appropriate diagnostic information to guide correct treatment selection. In particular, 54% (7/13) low-risk B-ALL and potentially therapy-changing high-hyperdiploid and *DUX4*r subtypes were identified with asWGS after being missed by conventional testing. Five cases with disease-defining Ph-like fusions were newly identified, including *ETV6::NTRK*, two *IGH::EPOR* cases, and two *PAX5::JAK2* cases. Among “other” AML cases on clinical testing, NASVar resolved the subtype in 67% (10/15), including *KMT2A*, *NUP98*r, *RUNX1*::*RUNX1T, NPM1*, and *CEPBA*, each with therapeutic implications.

#### Pharmacogenomics

After excluding the PDX specimens, *TPMT* and *NUDT15* were identified in 4% (19/431) and 7% (31/431) of cases, respectively. PMC had no cases of *NUDT15* variants, 6/124 (5%) with heterozygous *TPMT* variants, and 4/124 (3%) with homozygous TPMT variants. In contrast, at TMC-K and IHHN combined, 14% (29/201) of cases had *NUDT15* variations (including one homozygous case) and only 2% (4/201) of cases had *TPMT* variants.

### Implementation Outcomes

#### Feasibility

All four hospitals successfully validated asWGS, demonstrating feasibility across different contexts.

#### Fidelity

Two minor adaptations to the LDT framework and validation were identified. First, while UNC and PMC conducted limit-of-detection testing, UNC conducted *in vitro* testing aligned with CAP requirements, while PMC added an additional *in silico* evaluation based on center preferences. IHHN and TMC-K both included the testing completed and shared by UNC and PMC as part of their validation and conducted limited and no LOD testing, respectively, as a cost saving measure. Second, while UNC and PMC elected to continue singleplexing, TMC-K and Indus elected to implement a three-plex approach as a cost-containment strategy and justified by their validation findings.

#### Cost

The results of the microcosting analysis are presented in Figure 6. Based on United States pricing, costs ranged from $947/sample for a single case to $343/sample when three cases were multiplexed (**Figure 6A**). Flow cells were the main driver of costs. As expected, the cost/sample declined rapidly with multiplexing; however, the impact of multiplexing diminished as more samples/flow cell were used. Across centers, the difference in total cost using lowest and highest prices is 31.3% assuming pooled three-plexing (**Figure 6B**). A resequencing rate of 15% was still acceptable below the willingness to pay threshold (**Figure 6C**).

**Figure 6:**
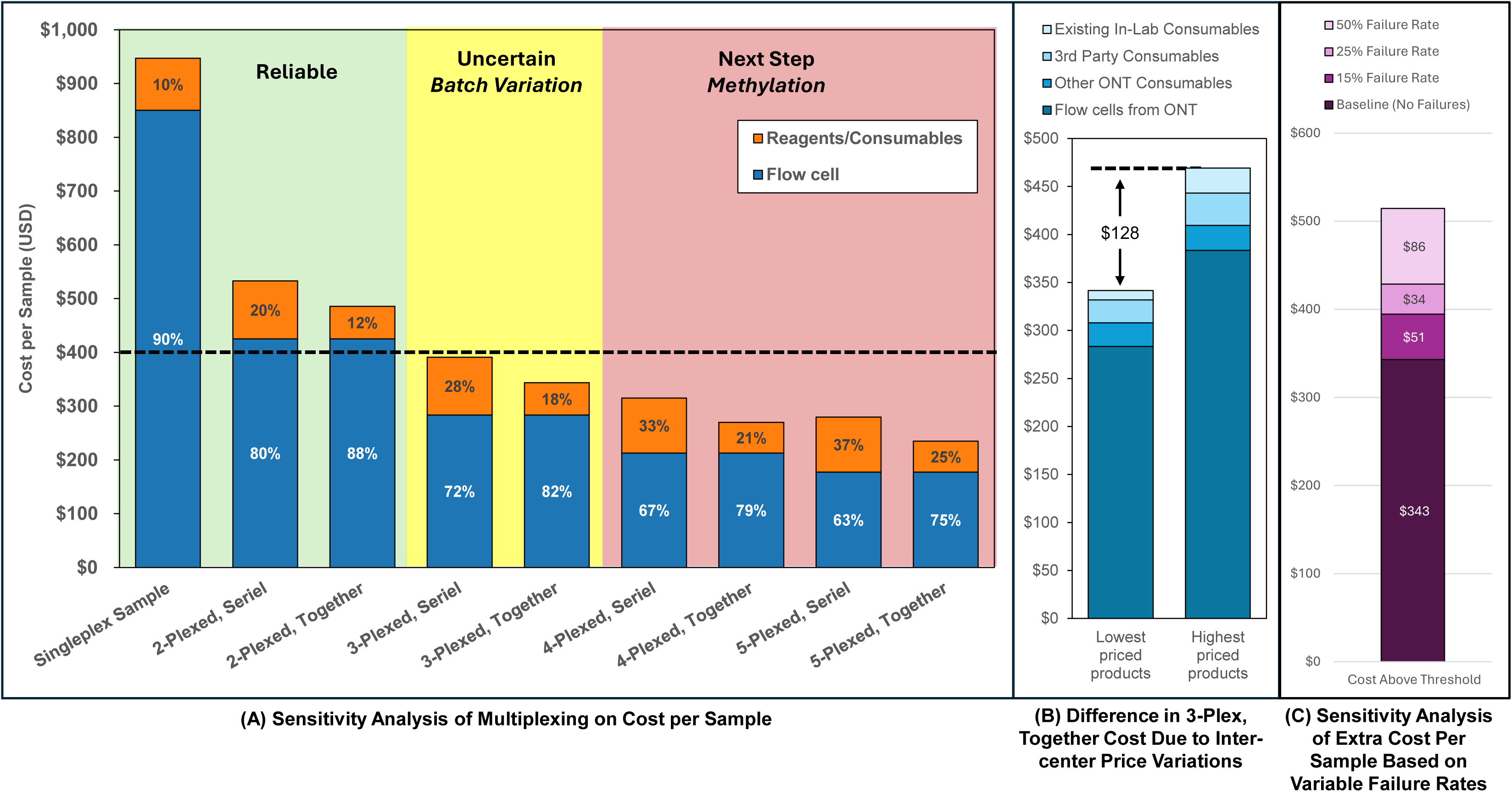
Microcosting Evaluation of Consumables and Reagents Required for Adaptive Sampling DNA Extraction, Library Preparation and Sequencing

#### Appropriateness

Each center is moving forward with clinical use of the assay. UNC is implementing within their clinical lab as a standard diagnostic for pediatric leukemia and adult B-ALL. Similarly, IHHN has elected to use the assay as standard of care for all new B-lineage acute leukemia patients. Given the existing NGS platforms in place, PMC will implement incrementally, focusing on leukemia subtypes where diagnosis is difficult or where rapid diagnosis is useful for clinical decision-making (e.g., *GLIS2*r in AML or *KMT2A* fusions in infant ALL). TMC-K is completing local regulatory requirements and finalizing a long-term reimbursement model prior to presenting a plan to hospital leadership.

## Discussion

Global access to clinical genomics remains a major impediment to rapid and equitable improvements in the treatment of childhood leukemias. To our knowledge, this study represents the first multicenter hybrid effectiveness and implementation study of a comprehensive NGS cancer diagnostic assay across diverse global contexts. Unlike traditional single-center assay development methods, we rapidly generated a sufficiently large dataset to demonstrate that clinically actionable information obtained using ONT-based asWGS is sufficient to accurately risk-stratify patients and inform dose adjustments for nearly all pediatric leukemia cases by pooling accuracy, precision, and replicability outcomes across multiple sites. At the same time, since each lab validated the assay using a common but independent LDT approach, we substantially reduced the lead time for hospitals to translate the findings from assay development to clinical return of results.

The direct benefit of asWGS over other diagnostic strategies is the added clinical utility across all diagnostic environments. Today, turnaround times in advanced pathology labs with robust conventional diagnostics and integrated NGS workflows varies from days to weeks; a limitation that impacts clinical decision-making. For example, identifying *DUX4*r B-ALL is difficult with conventional approaches and often requires weeks with NGS. asWGS can confirm the diagnosis within hours and provides a clinical research pathway to de-intensify highly toxic induction therapy for these patients.(25) Similarly, rapidly distinguishing between *PDGFRB* or *CSF1R* alterations in ALL can inform early selection of appropriate tyrosine kinase inhibitors (26). For high-risk AML subgroups, workflows using asWGS can identify chemotherapy refractory subtypes such as *GLIS2*r and *NUP98*r, dose-altering pharmacogenomics and near real-time polygenic single nucleotide variants to facilitate risk score calculation. With this information in hand, clinicians and research investigators would have the means to realize personalized therapeutic decision-making and the design of innovative frontline clinical trials, long-sought-after goals that are not feasible using currently standard diagnostic modalities (27–32).

From an equity standpoint, most patients and providers lack access to contemporary molecular diagnostics and routine NGS services both domestically in the United States and worldwide. Efforts to expand current evidence-based conventional diagnostic test for all children with leukemia at scale is complicated by the need for multiple tests (**Figure 1**), associated supply chain challenges, training across multiple assays, and limited comprehensiveness.(33) Meanwhile, the promise of NGS based on short-read WGS or RNAseq to simplify the leukemia-specific diagnostic cascade has been limited by similar challenges plus high costs and significant computational complexity (16,34). Additionally, WGS and WTS are complementary tests, requiring laboratories to run both assays orthogonally to achieve a comprehensive diagnosis.(35) Comparatively, asWGS across all four centers was completed using a simplified bioinformatics and diagnostic supply chain, together with relatively low consumable costs, thus providing strong evidence that expanding access to ONT-based asWGS is a viable and scalable technical solution toward the goal of democratizing equitable clinical NGS cancer diagnostics worldwide.(36) Moreover, our nested subanalysis analysis of the B-lineage “other” subgroup highlights the added clinical value of asWGS to provide simultaneous DNA alteration calling and methylation profiling from a single multi-omic assay, thus providing comprehensive integrated information resulting in cost-savings and obviating the need for multiple, overlapping tests (**Figure 1**).

At a health systems level, the successful treatment of a child with cancer requires an integrated and functioning value chain, including access to diagnostic capacity and appropriate therapies. Unfortunately, breaks in this value chain in low- and middle-income countries have resulted in estimated 5-year net survival rates of 20 to 30%, well below the >80% observed in high-income countries.(1) To address this disparity, the World Health Organization and St. Jude launched the Global Initiative for Childhood Cancer in 2018 with the goal of achieving 60% survival for children with cancer by 2030. Global commitment to this target was further reinforced in 2025 at the 4th High-Level Meeting on Non-Communicable Diseases, where member states endorsed accelerated action to reduce non-communicable diseases, with specific reference to the GICC 60% survival target.(37) While these calls for action have led to interventions expanding access to cytotoxic therapies and radiotherapy through programs such as the GPACCM, the IAEA–St. Jude Rays of Hope for Childhood Cancer initiative, and industry-led efforts such as Act for Children, there has been limited parallel progress in expanding access to comprehensive, accurate, and cost-effective diagnostic services.(38,39) Against this pessimistic backdrop, our data provide a new roadmap for equitable diagnostic scale-up worldwide. Next step efforts should focus on further driving down costs and improving supply chains through innovative procurement strategies such as service-level agreements, pooled purchasing, and increased price transparency mechanisms.

While our type-two hybrid effectiveness-implementation validation study results appear promising, several limitations were identified. First, the aneuploid and copy number classification currently requires >50% blasts in the specimen. While this may limit rare cases, almost all cases of ALL present with marrow blast percentage above this threshold. Second, although the assay performs well for variants currently included within NASvar, known rare variants were missed from automated calling. Fortunately, manual review of the asWGS matched the lesions observed from short-read testing and in real-world situations, concomitant DNA methylation analysis could have signaled a focused review. These findings highlight significant headroom for improvements in future NASvar versions when calling uncommon but established variants. Relatedly, the assay is being optimized to account for integration of methylation on top of variant calling. As a next step, we expect individual hospitals worldwide will need to consider the value of information costs and trade-offs between accepting high-probability results from shallow but rapid methylation profiling as a diagnostic tool for clinical determination versus requiring full structural variant calling. As additional data is generated, we anticipate the need for a resource-adaptable value and ethics framework to guide hospitals with lower willingness-to-pay thresholds in decision-making.(40) Finally, the microcosting analysis only included reagent and consumable costs and did not factor in labor or capital expenses. A key benefit for ONT is that historically the capital equipment prices have been substantially lower compared to other industry peers. Recent changes in the hardware lineup at ONT, however, threaten the long-term viability of scale-up. Although future work should include labor expense, the option to de-implement existing conventional pathology assays suggests labor is a sunk cost, with likely no additional human resource expenses beyond retraining required.

In conclusion, too many children die due to misdiagnosis or lack of diagnostic capacity. ONT asWGS is a realistic solution to address worldwide equity gaps in access to comprehensive clinical genomics. Moreover, and agnostic of technique or chemistry, our hybrid validation study design, driven by principles of seeking consensus, establishing standards and identifying value from ideation through implementation, provides a novel case-study to reduce known diagnostic implementation gaps and improve the quality of care.(41)

## Data Availability

All data produced in the present study are available upon reasonable request to the authors

